# Can infrared camera images be used for screening in Adolescent Idiopathic Scoliosis?

**DOI:** 10.1101/2021.10.16.21265088

**Authors:** Lam Oi Ka Natalie, James Cheng Peng, Teng Zhang, Joanne Yip, Queenie Fok, Maple Lee, Charlotte Wong, Kenny Kwan, Kenneth MC Cheung

## Abstract

**Introduction/Background context:** Adolescent idiopathic scoliosis (AIS) is the most common type of scoliosis, and affects up to 4% of adolescents in early stages^1^. The deformity can develop during any of the rapid periods of growth in children^2^, and the time of pubertal growth spurt also plays a role in spinal curve progression^3^. Hence it is crucial to detect the disease early, to provide timely intervention.

Detection of scoliosis when it is mild or before the growth spurt can be conducted via various screening methods. Adam’s forward bend test (FBT) and scoliometer measurement of the angle of trunk rotation (ATR) are commonly used^4^, to observe lateral bending and rotation of the spine, causing a visible rib hump. Moiré topography can also be used, but is reserved for second tier due to some degree of ambiguity^5^. X-rays (XR) remain the best way to diagnose scoliosis, as it provides a clear image of the spine and allows measurement of Cobb angle; however it has risks associated including requirement of the use of ionising radiation^6^.

Infrared (IR) thermography can be used to measure surface temperature and is performed with an IR camera. The temperature distribution and data matrix can be visualised into a thermal map, which has previously been studied and associated with the thermal asymmetry in paraspinal muscles^7^, as well as significant temperature differences between the convex and concave side of the spinal curvature for idiopathic scoliotic patients^8^. We hypothesize that such asymmetry and temperature differences may produce a detectable pattern on IR thermography, which would prompt further confirmatory investigations to reach a fast and non-radiation screening of AIS.

*Aims and Objectives:* For the purposes of this study, the hypothesised thermal pattern detected by IR thermography will be compared to the spinal deformities observed on XR, the ground truth as labelled by specialists. This will allow us to establish a paired dataset comparing spine visualisation between the two imaging modalities. Thus, the compared dataset would enable the generation of a machine learning model capable of automated classification of the deformity severities. The predictive accuracy of the model will be evaluated by the specialists’ assessment results. The aim of the project is to use IR to predict the severity of the scoliotic curve, opening up the possibility of using IR thermography with no radiation exposure and fully automated analytical pipeline for the screening of AIS.

## Methods and Materials

### Dataset and preprocessing

#### Subjects

Patients diagnosed with AIS at a specialist clinic from May 2018 to August 2019 who met the Scoliosis Research Society (SRS) Brace Manual criteria^9^ were enrolled in the study^10^. The inclusion criteria are: male or female at age between 10-15 years old at the time of consent provided, skeletal immaturity defined by Risser grade^11^(amount of ossification and eventual fusion of the iliac apophysis) of 0-2 inclusively. For females, they must be either premenarchal or less than 1 year post-menarche. Exclusion criteria were no other diagnoses apart from AIS, disabilities or systemic illnesses and no prior treatment was received. Ethics was approved by the Institutional Review Board of the University of Hong Kong/ /Hospital Authority Hong Kong West Cluster (HKU/HA HKW IRB) – IRB Reference number: UW17-136.

#### IR Camera and Experimental Procedure

Having met the requirements, participants were then referred for infrared thermography at Hong Kong Polytechnic University^8^. Subjects were asked to rest for 20 minutes to allow for acclimatisation prior to taking the image. They were also asked to avoid caffeinated beverages and exercise 4 hours before^12^. After undressing, markers were placed on the subjects’ back, according to the landmarks recommended by the Society on Scoliosis Orthopaedic and Rehabilitation Treatment (SOSORT) guidelines^13^. The temperature of the room was controlled to 20 °C ± 2 °C and a relative humidity of 55%^14^. The IR thermal images were captured using a FLIR E33 camera (FOL-18 lens; 10,800 pixels) in a dark room, placed on a tripod 1.5m away from the patients. The camera has a thermal sensitivity of 0.07 °C and camera emissivity level of 0.98. This was conducted in accordance with the Helsinki Declaration of the World Medical Association Assembly, and ethics was approved by the Human Subjects Ethics Sub-committee (HSESC) of the Hong Kong Polytechnic University. All participants and/or their legal guardian provided written informed consent regarding the data for study.

#### Data Visualisation and Normalisation Methods

The visualisation of the IR images is implemented using the *OpenCV* package (Fig. 1). The images were then processed by a computer to generate a thermal matrix dataset of CSV files, each representing the thermal matrix of one patient’s back surface. They were then processed and standardised (Fig. 2) via normalisation methods, to improve the model performance in the severity prediction. This included removing the background temperature and adjusting the visualisation sensitivity.

**Figure 1:**
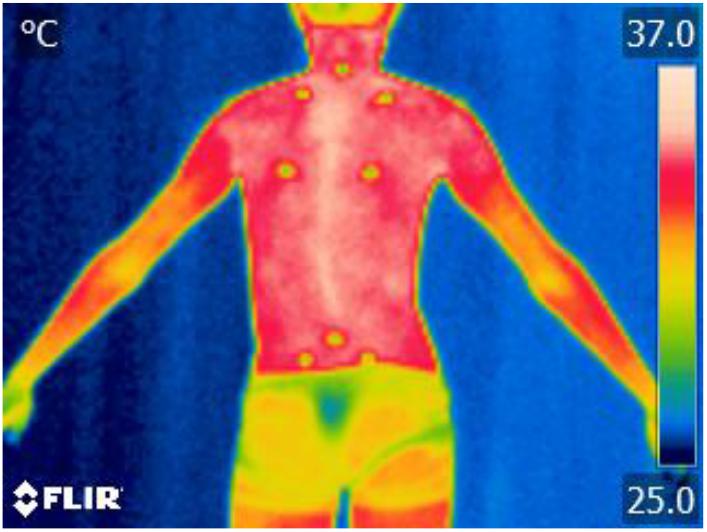
Original IR Thermal spine image

**Figure 2:**
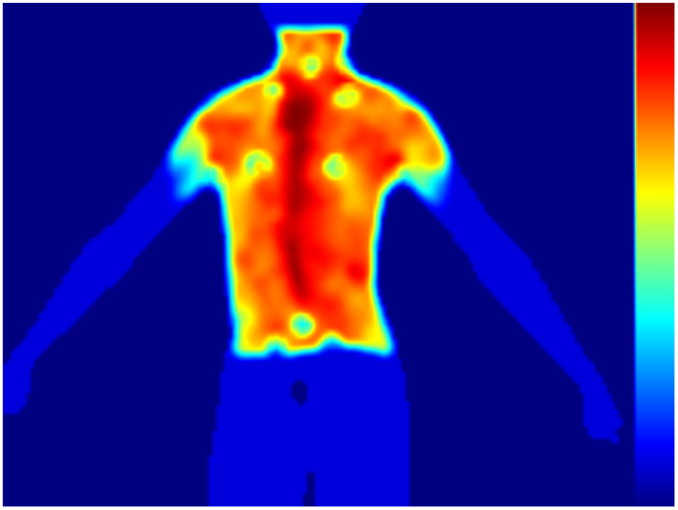
Standardised processed spine image

#### Generation of ground truth of the spinal deformities

PA radiographs were taken for all patients for diagnostic purposes of AIS, which also serves as a baseline image for the subjects’ spine (Fig. 3). Coronal cobb angles of the major and minor curve(s) were measured, and these were considered the ground truth of the structural curvature. Location, directionality and magnitude of the curves (major curve Cobb angles) were then compared to the pattern from the IR thermal matrix generated, creating a paired dataset.

**Figure 3:**
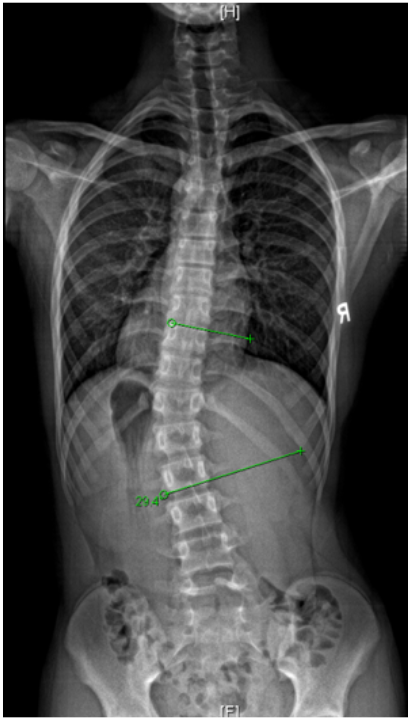
XR spine (ground truth)

#### Machine learning based severity classification

Traditional machine learning methods which are not based on artificial neural networks, were opted for in this study. They do not require a large amount of data sample for model fitting, which is necessary for deep learning methods. Three ensemble learning models were used, namely XGBoost^15^, AdaBoost^16^ and Random Forest^17^. One common feature of these three models is that they train a set of decision trees, and subsequently ensemble the results given by each tree in order to increase the robustness. In general, ensemble methods help partially overcome three problems which arise from non-ensembled methods^18^. The first is the statistical problem which always occurs when the space of hypothesis is too large with regards to the amount of training data. In this case, there may be multiple hypotheses that give the same accuracy. Ensemble methods mitigate the risks of choosing an inappropriate hypothesis by taking the votes of many equally good classifiers. The second problem lies in computation. During the training of one classifier, due to the non-convexity of the objective function, the model parameters may end up with the local optima. A combination of multiple different local optima would mitigate the risk of choosing the wrong local optima. The last problem is representation. This arises when there does not exist a hypothesis in the hypothesis space that can capture the true reality. To some extent, taking a combination of hypotheses expands the set of functions that can be presented. Hence, we employed the tree-based ensemble learning methods in this study, which have strong robustness.

Decision trees are a class of non-parametric supervised learning methods consisting of tree nodes and branches. They map an input of vector form to a categorical (for classification tasks) or scalar (for regression tasks) output. At each non-leaf node, one element of the input (attribute), is considered and a decision threshold with respect to this attribute determines which branch a data sample goes to. One common algorithm for attribute selection is the Gini index^19^. Suppose p_i_ is the probability that an arbitrary data point in the dataset D belongs to class C_i,_ then the Gini index for D is calculated by:

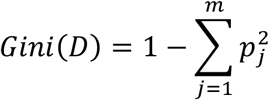

where m is the number of classes in the dataset. Suppose we partition the datatset D based on attribute A into two subsets D_1_, D_2_, in other words, the data points in D_1_ have value in attribute A smaller than a threshold a_k_ while the data points in D_2_ have value in attribute A larger than a_k,_ then the Gini index based on attribute A and the threshold a_k_ is:

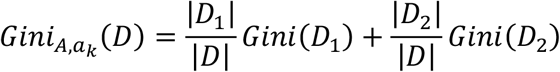

In this case, we would select attribute A and splitting threshold a_k_ with the smallest Gini index. Note that we need to enumerate all the possible splitting points for each un-used attribute.

When it comes to ensemble learning, multiple trees are trained by subsets of attributes and samples, and restrictions on tree depth are imposed to prevent overfitting. In this study, we reshaped each thermal image with size 120*160 into a row vector with length 19200 before passing into the ensemble models.

The dataset from the paired imaging modalities, was subsequently fed into the three machine learning models, in order to teach each model to classify images based on the severity of the spinal curvature. Train-test split procedure was performed for evaluation. Split percentage of the sample was train: 70%; test: 30%. The accuracy of the model was assessed by comparing the classification of the severity of the curve based on IR thermography, to the true severity based on Cobb angle from XR. Correct identification into the class was considered a positive result.

#### Statistical assessments

4 classes were used to differentiate between severity of curves based on coronal Cobb angle: Group 0 < 20°, Group 1 20°-30°, Group 2 31°-40°, Group 3 > 40°. Resampling techniques were employed to artificially rebalance the dataset to avoid under-sampling the majority class or over-sampling the minority class. The minority class was oversampled to augment the original dataset using SMOTE: Synthetic Minority Over-sampling Technique ^20^, without sacrificing the existing numbers. Evaluation metrics were selected for comparison between machine learning performances include precision/sensitivity, recall/ positive predictive value, f1-score, weighted averages, macro averages and accuracy. Furthermore, cases where the spine could not be clearly mapped out from the IR image (eg. Fig 4) were manually excluded, generating a filtered sample with outliers excluded. The model training was carried out using both original and filtered data. Student’s *t* test (p<0.05) was used to compare performance measures.

**Figure 4:**
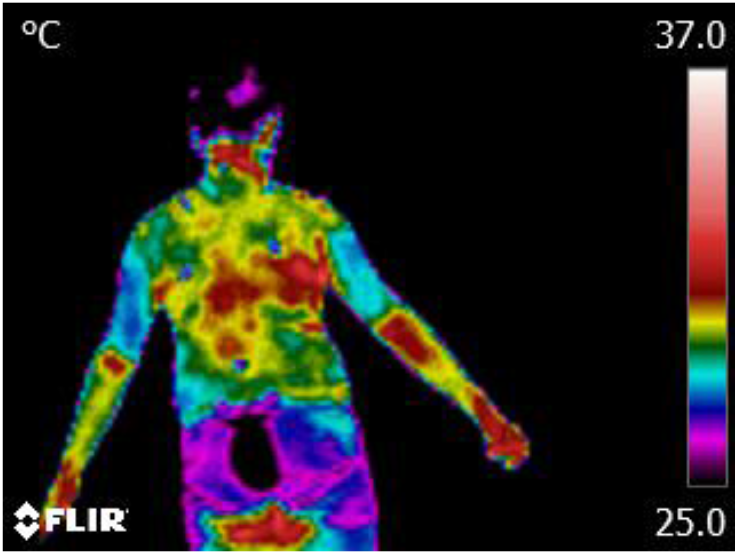
IR Thermal spine image where spine is not identifiable

## Results

### Subjects

82 participants (18 males and 64 females) were recruited for the study (Fig. 5).

**Figure 5.**
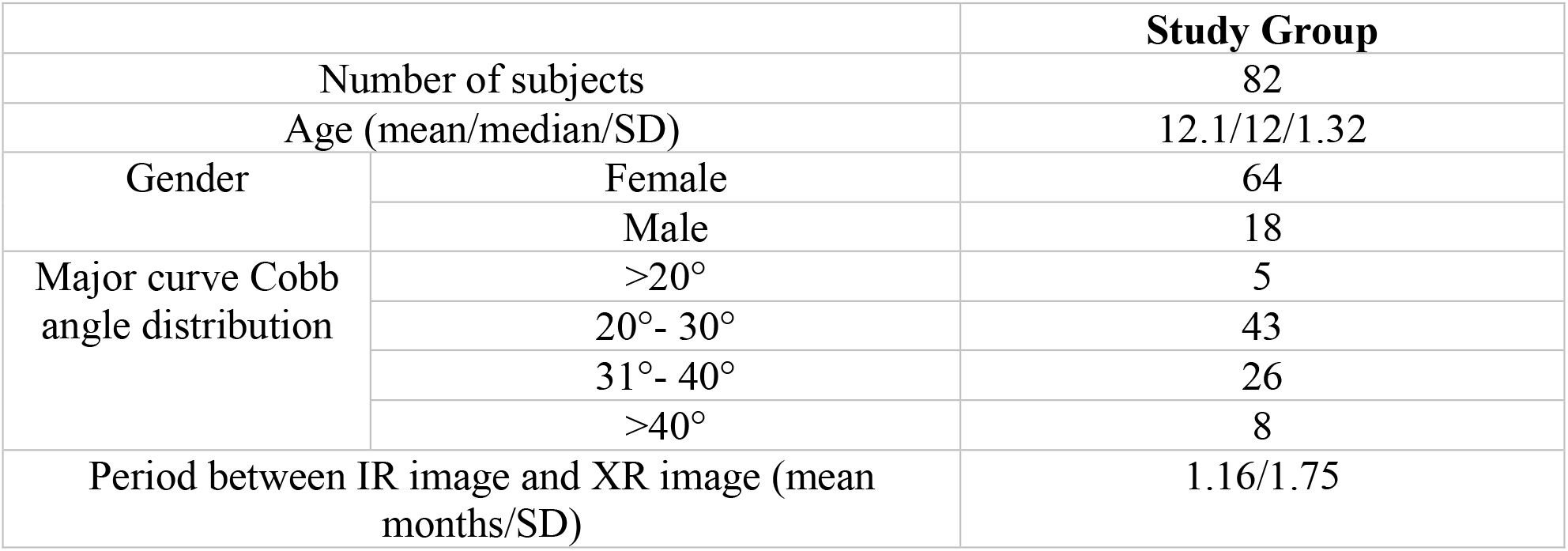
Baseline characteristics of study group

The severity of AIS of the sample group was imbalanced (Fig 6) and not normally distributed (Fig 7). Hence the clinical gold standard classification of severity of spinal curvatures (mild < 20°, moderate 20°-40°, severe > 40°) was not used, as the accuracy of the model severity prediction would be subject to frequency bias. Furthermore, 15 cases were excluded to produce a filtered set of a total of 67 curves.

**Fig 6:**
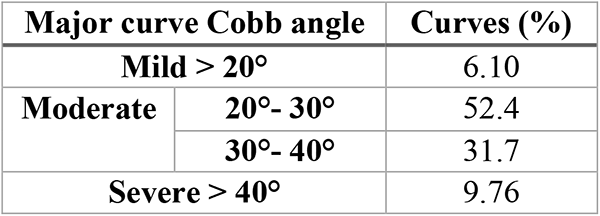
Distribution of Cobb angles in each class

**Figure 7:**
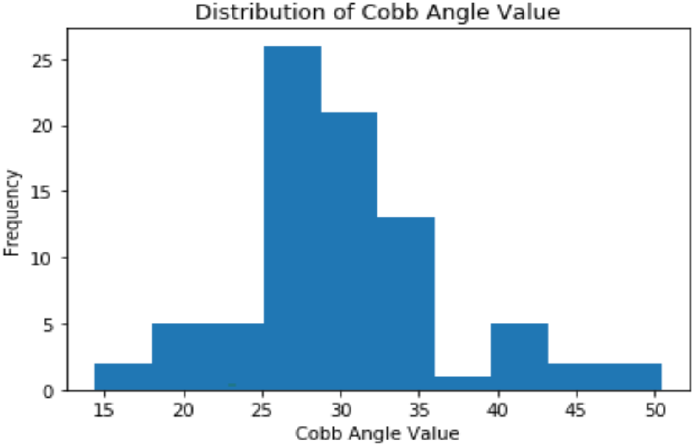
Cobb angles distribution.

### Data Visualisation

All pixel values were extracted and plotted, summing to a total of 1,574,400. The range of all pixel values were from 16.59 to 41.85. The distribution is positively skewed, where most of the pixels are of value between 20 and 30 (Fig. 8). There was also a large variation in environment temperature (20°+/-6°) across all original images, which was standardised after normalisation.

**Figure 8:**
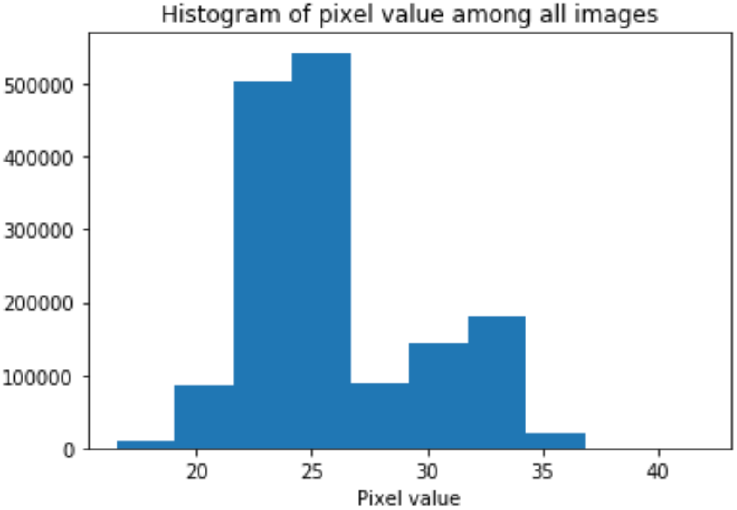
Image Pixel values. The X − axis represents the pixel value and the Y − axis represents the number of pixels.

### Result of severity prediction for the 3 machine learning models

Overall out of the 3 machine learning models, XGBoost and Random Forest performed better in all parameters than AdaBoost. After resampling, all performance metrics (precision, recall, f2-score, accuracy) improved statistically significantly for the two models, for both original (n=82) and filtered data (n=67). The accuracy for prediction of the severity of the curve for the original data were XGBoost = 0.83, Random Forest= 0.81. All weighted parameters (precision, recall, f1-score) also reached >=0.80. However, the accuracy for AdaBoost was 0.66, and none of the other parameters reached 0.70 in either original or filtered data. In terms of filtering the dataset, there was no statistically significant improvement in overall accuracy or classwise performance for all 3 models.

In terms of class-wise performance, the precision, recall and f1-score were mostly highest in groups 0 and 3. Precision was 0.88 and 1.00 for each group respectively for both XGBoost and Random Forest. This implies that the model was strongest for the mildest and most severe curves, which may not be reliable and could be attributed to oversampling of the minority class. XGBoost mildly outperforms Random Forest for all classes, including precision of group 1 (0.77; 0.71) and group 2 (0.69; 0.67). However the weighted averages of precision and recall for the two models were not significantly different. Fig 9-11 shows the accuracy and weighted averages for both data before and after resampling, for both datasets, for the different models.

**Fig 9.**
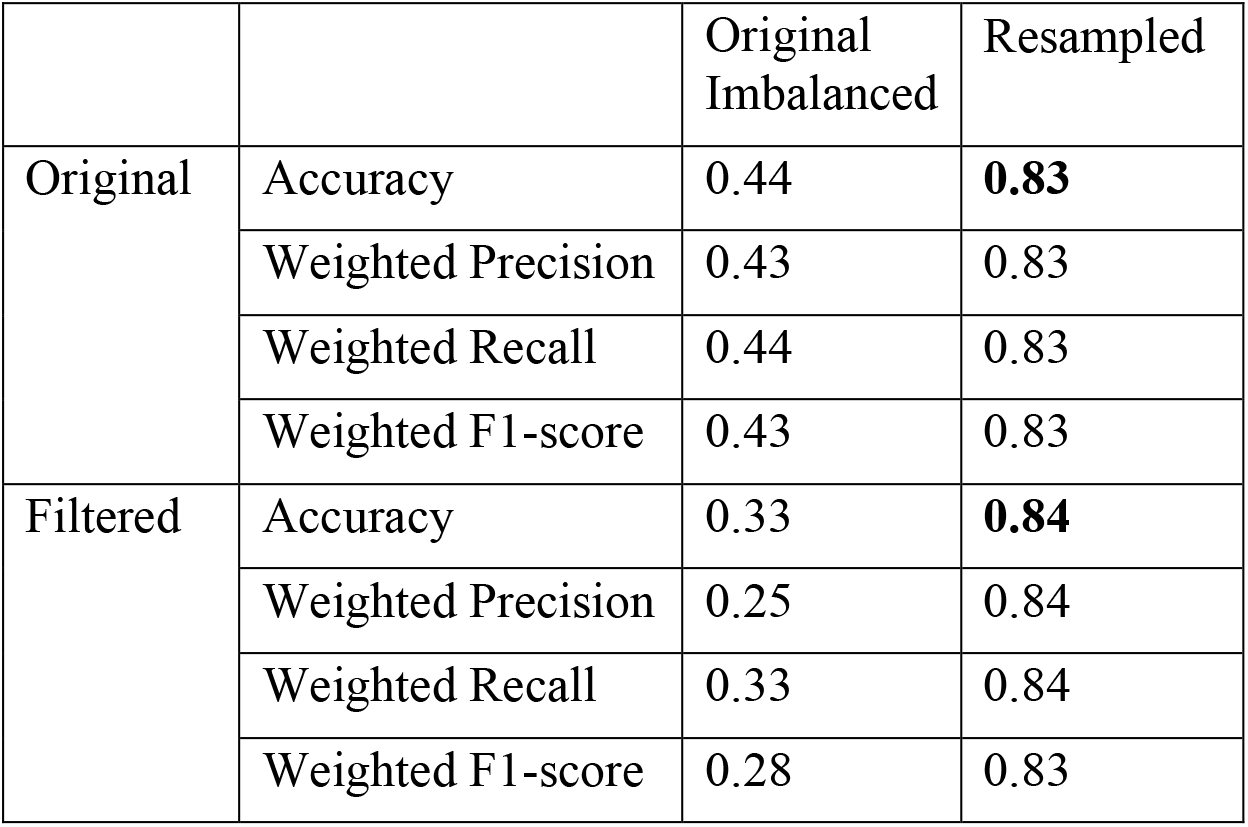
Accuracy and weighted averages for XG Boost

**Fig 10.**
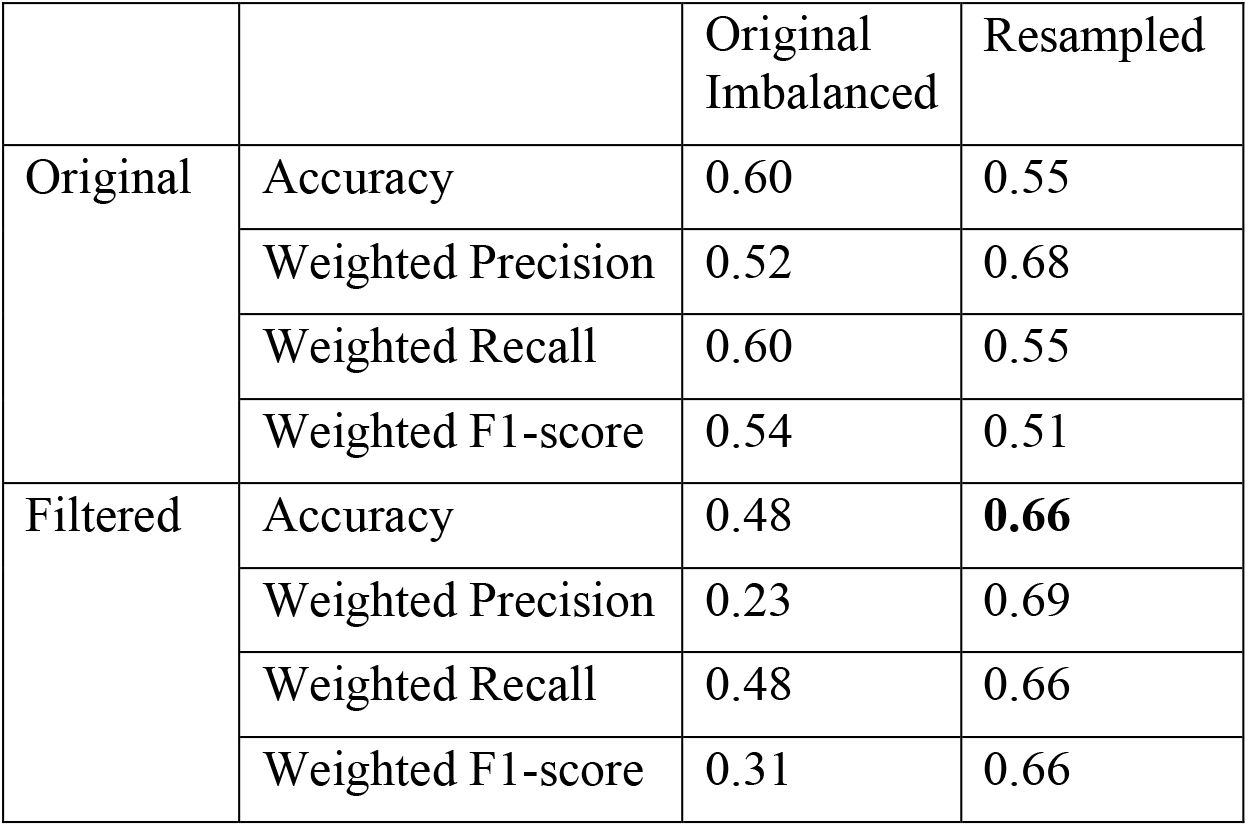
Accuracy and weighted averages for AdaBoost

**Fig 11.**
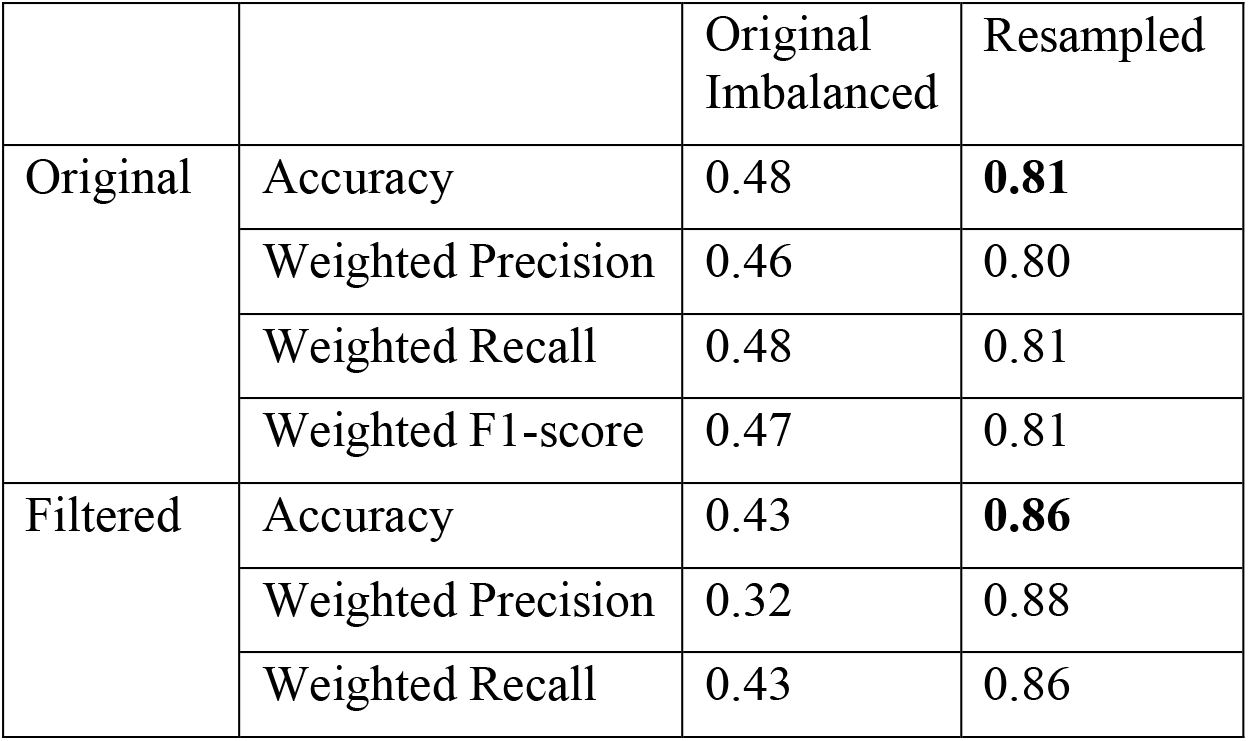
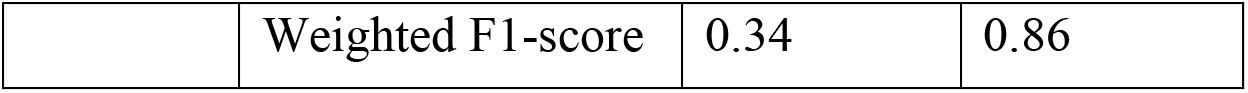
Accuracy and weighted averages for AdaBoost

### XG Boost

### AdaBoost

### Random Forest

### Raw Data with class wise performance

#### 1. Original Data

##### Imbalanced Data

*XGBoost*
|  | precision | recall | f1-score | support |
| --- | --- | --- | --- | --- |
| 0 | 0.00 | 0.00 | 0.00 | 1 |
| 1 | 0.59 | 0.62 | 0.61 | 16 |
| 2 | 0.20 | 0.14 | 0.17 | 7 |
| 3 | 0.00 | 0.00 | 0.00 | 1 |
| accuracy |  |  | 0.44 | 25 |
| macro avg | 0.20 | 0.19 | 0.19 | 25 |
| weighted avg | 0.43 | 0.44 | 0.43 | 25 |

*AdaBoost*
|  | precision | recall | f1-score | support |
| --- | --- | --- | --- | --- |
| 0 | 0.00 | 0.00 | 0.00 | 1 |
| 1 | 0.67 | 0.88 | 0.76 | 16 |
| 2 | 0.33 | 0.14 | 0.20 | 7 |
| 3 | 0.00 | 0.00 | 0.00 | 1 |
| accuracy |  |  | 0.60 | 25 |
| macro avg | 0.25 | 0.25 | 0.24 | 25 |
| weighted avg | 0.52 | 0.60 | 0.54 | 25 |

*Random Forest*
|  | precision | recall | f1-score | support |
| --- | --- | --- | --- | --- |
| 0 | 0.00 | 0.00 | 0.00 | 1 |
| 1 | 0.62 | 0.62 | 0.62 | 16 |
| 2 | 0.22 | 0.29 | 0.25 | 7 |
| 3 | 0.00 | 0.00 | 0.00 | 1 |
| accuracy |  |  | 0.48 | 25 |
| macro avg | 0.21 | 0.23 | 0.22 | 25 |
| weighted avg | 0.46 | 0.48 | 0.47 | 25 |

##### Resampled Data

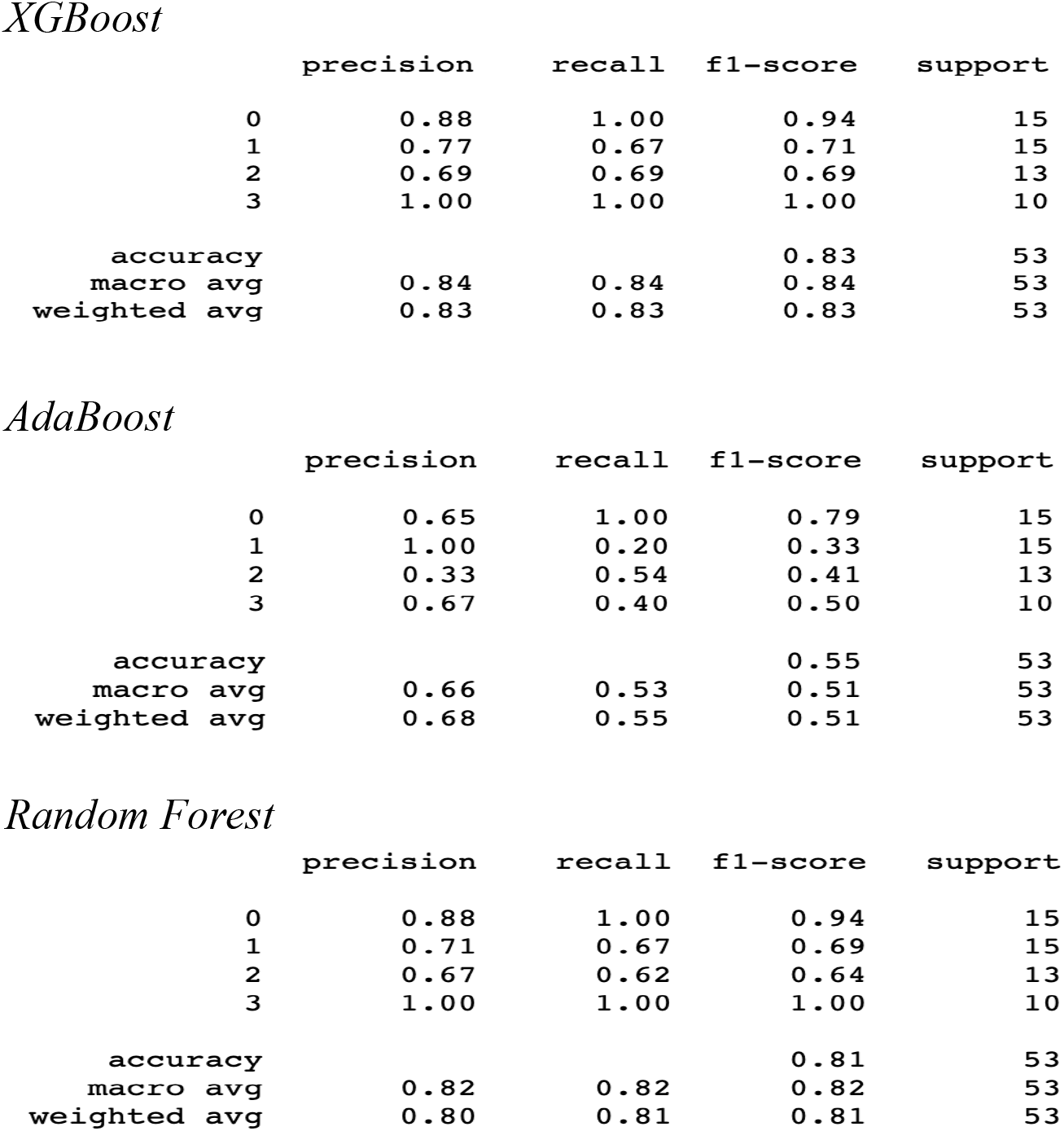

#### Filtered Data

##### Imbalanced Data

*XGBoost*
|  | precision | recall | f1-score | support |
| --- | --- | --- | --- | --- |
| 0 | 0.00 | 0.00 | 0.00 | 1 |
| 1 | 0.38 | 0.60 | 0.46 | 10 |
| 2 | 0.20 | 0.14 | 0.17 | 7 |
| 3 | 0.00 | 0.00 | 0.00 | 3 |
| accuracy |  |  | 0.33 | 21 |
| macro avg | 0.14 | 0.19 | 0.16 | 21 |
| weighted avg | 0.25 | 0.33 | 0.28 | 21 |

*AdaBoost*
|  | precision | recall | f1-score | support |
| --- | --- | --- | --- | --- |
| 0 | 0.00 | 0.00 | 0.00 | 1 |
| 1 | 0.48 | 1.00 | 0.65 | 10 |
| 2 | 0.00 | 0.00 | 0.00 | 7 |
| 3 | 0.00 | 0.00 | 0.00 | 3 |
| accuracy |  |  | 0.48 | 21 |
| macro avg | 0.12 | 0.25 | 0.16 | 21 |
| weighted avg | 0.23 | 0.48 | 0.31 | 21 |

*Random Forest*
|  | precision | recall | f1-score | support |
| --- | --- | --- | --- | --- |
| 0 | 0.00 | 0.00 | 0.00 | 1 |
| 1 | 0.44 | 0.80 | 0.57 | 10 |
| 2 | 0.33 | 0.14 | 0.20 | 7 |
| 3 | 0.00 | 0.00 | 0.00 | 3 |
| accuracy |  |  | 0.43 | 21 |
| macro avg | 0.19 | 0.24 | 0.19 | 21 |
| weighted avg | 0.32 | 0.43 | 0.34 | 21 |

#### Resampled Data

Resampled Data XGBoost
|  | precision | recall | f1-score | support |
| --- | --- | --- | --- | --- |
| 0 | 0.83 | 1.00 | 0.91 | 15 |
| 1 | 0.82 | 0.69 | 0.75 | 13 |
| 2 | 0.67 | 0.60 | 0.63 | 10 |
| 3 | 1.00 | 1.00 | 1.00 | 12 |
| accuracy |  |  | 0.84 | 50 |
| macro avg | 0.83 | 0.82 | 0.82 | 50 |
| weighted avg | 0.84 | 0.84 | 0.83 | 50 |

AdaBoost
|  | precision | recall | f1-score | support |
| --- | --- | --- | --- | --- |
| 0 | 0.90 | 0.60 | 0.72 | 15 |
| 1 | 0.44 | 0.54 | 0.48 | 13 |
| 2 | 0.50 | 0.50 | 0.50 | 10 |
| 3 | 0.86 | 1.00 | 0.92 | 12 |
| accuracy |  |  | 0.66 | 50 |
| macro avg | 0.67 | 0.66 | 0.66 | 50 |
| weighted avg | 0.69 | 0.66 | 0.66 | 50 |

Random Forest
|  | precision | recall | f1-score | support |
| --- | --- | --- | --- | --- |
| 0 | 0.83 | 1.00 | 0.91 | 15 |
| 1 | 1.00 | 0.62 | 0.76 | 13 |
| 2 | 0.67 | 0.80 | 0.73 | 10 |
| 3 | 1.00 | 1.00 | 1.00 | 12 |
| accuracy |  |  | 0.86 | 50 |
| macro avg | 0.88 | 0.85 | 0.85 | 50 |
| weighted avg | 0.88 | 0.86 | 0.86 | 50 |

## Discussion

The current screening process involves several different techniques and stages, which may be cumbersome and take a long time to reach a diagnosis. This paper sees to investigate whether the use of IR thermography would reduce the multi-step process into one single step, thereby increasing the efficiency of screening and reduce the amount of radiation patients are exposed to during repeated follow-ups. The integration of the two modalities may be complimentary and provide a more holistic overview as well as complete picture about the spine and back muscles.

The visualization functionality of the paired dataset aimed to provide a simple user-interface (UI), in which the user can click on one button to select the CSV file containing the thermal matrix from the computer, as well as the thermal image side by side. The UI is implemented by the *tkinter* library in Python. This allows both the original and processed thermal images to be compared to the ground truth XR. However, it was unable for the two programs to capture the spatial information stored in the image, and rather a flattened vector was required. This could be developed using other convolutional kernels, to record the specific gestures and actual images itself.

In terms of classification of the severity of the curve based on thermography, the overall performance for XGBoost and Random Forest shows the potential and feasibility to use traditional machine learning methods to classify the severity of scoliosis with fine-tuning from an IR thermal matrix. The precision, recall and f1-score were similar after resampling (all >=0.80) which therefore does not show particular weaknesses for false positives/negatives in these two models. It is evident that resampling plays an important role in our study in enhancing the robustness of the models. This technique was however employed due to imbalanced dataset. This may have resulted in over sampling of both Group 0 and 3 and caused overfitting in some situation. The precision, recall and f1-score were all 1.00 for both XGBoost and Random Forest after resampling. Interpretation of class-wise performance is therefore limited from our study. Recruitment of more samples was not possible due to the time constraint, in addition to the difficulty specifically recruiting only mild and severe cases. Nonetheless, with all parameters >=0.80 for two types of machine learning models, it shows the potential of integrating infrared thermography into the screening of AIS.

There were other limitations in the study due to the small sample size, including opting for traditional machine learning methods for analysis. More datasets can be used in the future, which would allow the implementation of deep convolutional neural network. Deep machine learning models may have the potential to outperform the traditional methods - XGBoost and Random Forest, and extract other patterns from the images that are not considered by human experts, potentially providing additional information on the spinal deformity. Another alternative is by adding attention to the network. However, as attention mechanisms were first applied in recurrent neural network for natural language processing which involves sequential data, how this can be retrieved from the thermal matrix and scoliotic patients need to be investigated further.

Secondly, there was difficulty in observing the spine on some thermography images due to various factors, such as clothing, background interferences and obesity of the patient (Fig. 12). Full exposure of the back with no clothing would allow a better and more complete visual field of the spine. Furthermore, the markers placed were also unnecessary for the purposes of this study. Omitting them would allow each scan to be conducted quicker, lowering the workload of the practitioner. Both improvements would also allow more data points into the thermal matrix.

**Figure 12:**
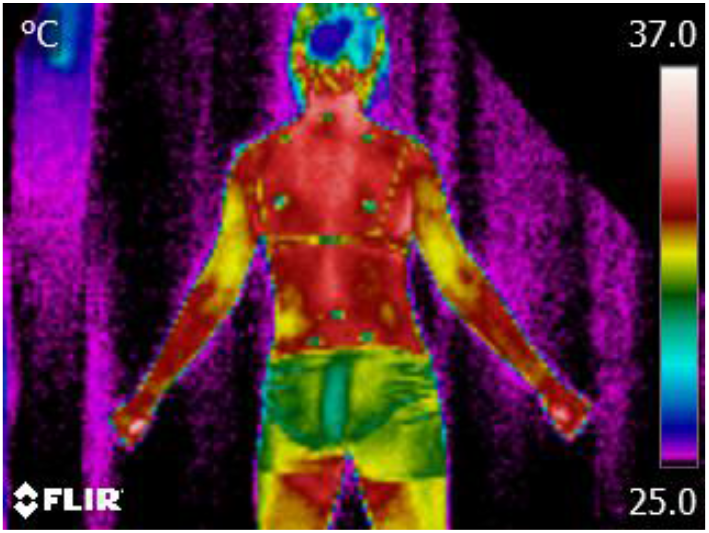
Thermal Image with patients wearing clothing and background interference

The BMI or waist:hip ratio of the patient can also be correlated with the spinal curve, to determine whether a BMI cut-off point should be set for IR thermography. It was notably more difficult to observe the spine on obese patients, as compared to thinner patients. They were thus excluded in the filtered dataset.

Additionally, the standing posture and arm position of patients should also be standardised. The subjects were not all facing directly ahead, with some turning to the side. The position of the arms were also held at a different angle away from the body in each patient (Fig 13.). The variation in posture may be accounted to difficulty in finding a focal point in a dark room, long waiting time between standing and actual photo taken and subject getting bored. Varied trunk rotational angles will affect the visualisation of the spine as well as detection of the temperature of the paraspinal muscles by IR. The arm position may also involve contraction of the back muscles, and hence affect surface temperature of the concerned region. As there is difficulty placing an entire frame in front of the 3D scanner for the patient’s back to lean against, a metal plate at the heel may be more suitable. Explicit instructions regarding the posture of the trunk and position of the arms should also be stated at the beginning prior to image taking.

**Figure 13:**
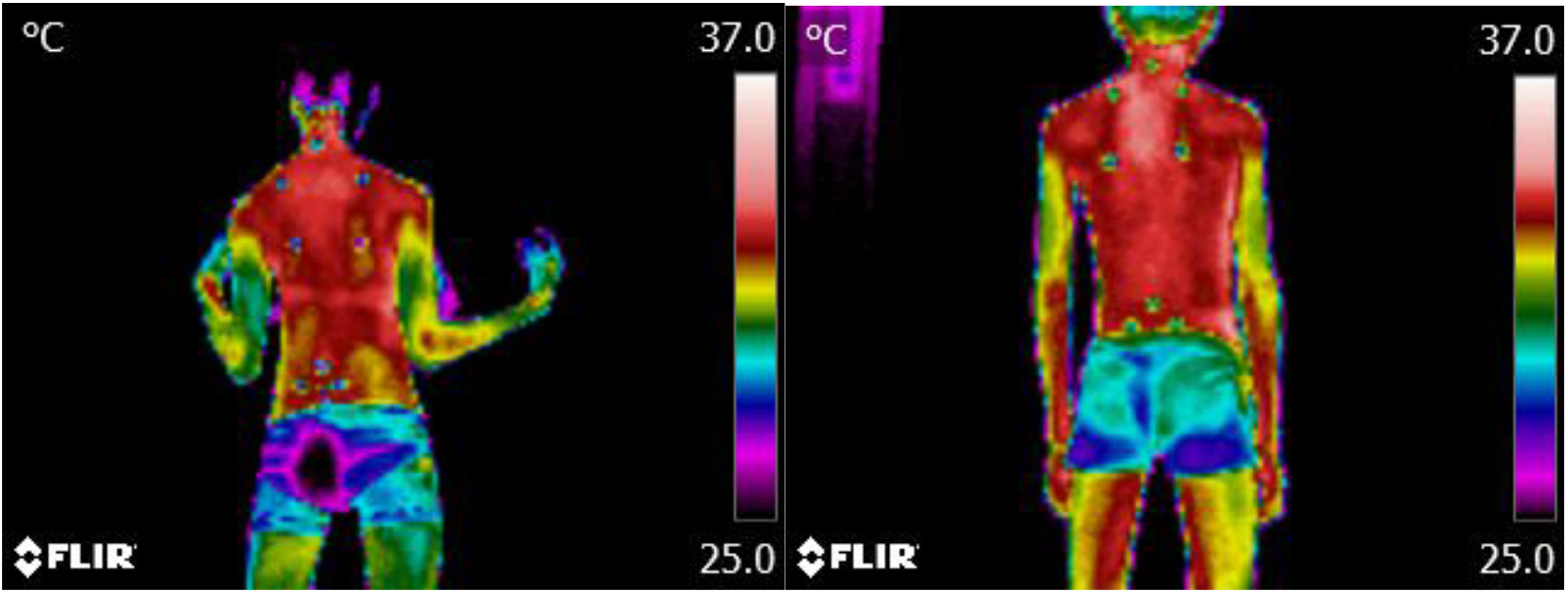
Thermal Image with patients holding arms at different positions

## Conclusion

The accuracy of prediction of severity of curves based on Cobb angles is > 0.80 for 2 machine learning types (XGBoost and Random Forest) after resampling. This shows promising potential for the use of infrared thermography to predict the severity of the scoliotic curve, and thus can be considered for screening of AIS, albeit its limitations in interpretation of class-wise performance. However, the actual feasibility and cost-effectiveness of integrating IR thermography into the current screening program has yet to be determined. A larger sample size would be required for validation, as there were only 10-15 cases in each class after resampling in this study. Furthermore, IR images of healthy non-scoliotic individuals would also be needed, in order to assess the usage and effectiveness of IR thermography for general screening of AIS.

## Data Availability

All data produced in the present study are available upon reasonable request to the authors

